# Comparative Efficacy and Safety of Antipsychotics for Parkinson’s Disease Psychosis: A Systematic Review and Network Meta-Analysis

**DOI:** 10.1101/2025.09.11.25335549

**Authors:** Sai Sahithi Reddy Atla, Praveen Gunasekaran, Shradha Pandurang Kakde, M Mithun, Abhinav Waghmode, Deepita Singh, Meghana Chennupati, Sumaiya Mehveen, M Abhinaya, Rakhshanda khan, Harshawardhan Dhanraj Ramteke

**Author notes:** Corresponding author Rakhshanda khan Ayaan institute of medical sciences, Moinabad, India.

## Abstract

**Background:** Psychosis affects over half of people with Parkinson’s disease (PD) over the disease course and severely worsens quality of life. Clinicians often face a trade-off between reducing dopaminergic therapies to control hallucinations/delusions and maintaining motor function. Multiple atypical antipsychotics are used, but their comparative efficacy and safety remain uncertain.

**Methods:** We conducted a prespecified systematic review and network meta-analysis (NMA) following PRISMA 2020 and registered in PROSPERO (CRD420251143957). PubMed, Embase, Scopus, and CENTRAL were searched to August 2025 without language restrictions. Randomized controlled trials evaluating atypical antipsychotics for PD psychosis were eligible. Two reviewers independently screened studies, extracted data, and assessed risk of bias (RoB 2); certainty was appraised with GRADE. Continuous outcomes (BPRS, CGI-S, UPDRS-II) were synthesized as mean differences; binary outcomes (adverse events, discontinuation, mortality, cardiovascular events) as risk ratios in a random-effects NMA, with placebo as the common comparator.

**Results:** A total of 22 trials with 2,047 participants (mean age 70.6 ± 12 years; 1,038 males and 997 females) were included, with a mean follow-up of 2.73 ± 1.0 months. Across treatment arms, 1,091 patients received active interventions and 1,036 placebo. The active groups included clozapine (n=139), olanzapine (n=111), pimavanserin (n=746), quetiapine (n=126), risperidone (n=5), and ziprasidone (n=8). None of the drugs demonstrated consistent or statistically significant superiority over placebo in reducing psychosis severity as measured by BPRS or CGI-S, although clozapine and quetiapine showed trends toward improvement and risperidone suggested possible benefit with very wide intervals due to small samples. Motor and daily living outcomes assessed with UPDRS-II revealed no significant changes across treatments, with pooled effects for clozapine, olanzapine, quetiapine, pimavanserin, risperidone, and ziprasidone all overlapping the null. Safety analyses indicated no meaningful increase in risk of PD worsening, insomnia, cardiovascular events, or mortality compared with placebo, with overall pooled risk ratios approximating unity and showing no heterogeneity. SUCRA rankings suggested risperidone and clozapine as potentially more efficacious on BPRS, olanzapine as higher on CGI-S, and ziprasidone on UPDRS-II, though none achieved robust statistical significance.

**Conclusions:** No antipsychotic showed clear superiority over placebo; clozapine and pimavanserin remain the most relevant options, but stronger evidence is needed.

## Introduction

Parkinson’s disease (PD) is a progressive neurodegenerative disorder primarily characterized by its motor manifestations—bradykinesia, rigidity, tremor, and postural instability [1]. However, beyond these hallmark features, non-motor symptoms significantly contribute to morbidity, disability, and diminished quality of life. Among these, **psychosis** is one of the most debilitating and prevalent complications [2]. Epidemiological data suggest that more than half of individuals with PD develop psychotic symptoms during the disease course, typically manifesting as hallucinations and delusions. With the global burden of PD projected to rise due to population aging, the absolute number of patients experiencing psychosis will inevitably increase, amplifying the clinical and societal impact of this condition.

The etiology of psychosis in PD is multifactorial. Neurodegenerative changes involving dopaminergic, serotonergic, and cholinergic pathways are implicated, but psychosis is also strongly associated with the pharmacologic therapies required to treat motor symptoms. Dopamine replacement therapies, while indispensable for mobility, can precipitate or exacerbate hallucinations and delusions [3]. Thus, the initial management strategy often involves reducing or discontinuing dopaminergic medications once psychosis emerges. This approach, however, creates a therapeutic paradox: alleviating psychosis by tapering dopaminergic drugs risks worsening parkinsonian motor disability, often to the point of impairing autonomy and daily function. This delicate balance between motor control and psychiatric stability constitutes one of the most formidable challenges in PD care.

Because of this clinical dilemma, **atypical antipsychotics (AAPs)** have been explored as potential solutions. These agents are considered due to their reduced propensity for extrapyramidal side effects compared with first-generation antipsychotics [4]. Yet, their use in PD psychosis has been fraught with limitations. Clozapine, one of the earliest AAPs studied, demonstrated robust efficacy in reducing psychotic symptoms without worsening motor function in randomized controlled trials [5]. Importantly, clozapine remains the only antipsychotic consistently supported by high-quality evidence in this population. Nevertheless, its widespread use has been hampered by a substantial risk of agranulocytosis and other hematologic complications, necessitating frequent blood monitoring and limiting its practicality in routine clinical practice. Observational studies, including analyses of prescribing data, confirm that clozapine remains underutilized despite its proven benefits [6].

Quetiapine, in contrast, has been more readily adopted in clinical practice, largely because it does not require intensive hematologic monitoring and is generally well tolerated with respect to motor function. However, the evidence base for its efficacy remains weak. Multiple RCTs and meta-analyses have failed to demonstrate consistent superiority of quetiapine over placebo, raising concerns that its widespread use may be driven more by convenience and perceived safety than by robust evidence of benefit. Other AAPs, such as olanzapine and risperidone, have been studied but are associated with unacceptable risks of motor worsening and are therefore not recommended in clinical guidelines.

A major shift in the therapeutic landscape occurred in 2016 with the approval of **pimavanserin**, the first agent specifically indicated for Parkinson’s disease psychosis [7]. Pimavanserin is a selective inverse agonist and antagonist of the 5-HT2A receptor, with negligible affinity for dopaminergic, histaminergic, or adrenergic receptors [8]. This unique pharmacologic profile was designed to provide antipsychotic efficacy while minimizing risks of extrapyramidal side effects. Phase III trials demonstrated meaningful reductions in psychosis severity with pimavanserin compared to placebo, coupled with an acceptable tolerability profile [9]. The introduction of pimavanserin has expanded therapeutic options and generated optimism among clinicians and patients. Nevertheless, long-term safety concerns—particularly related to cardiac effects and mortality—remain subjects of debate, and real-world effectiveness data are still emerging [10].

Despite these advances, **critical gaps persist** in the comparative evidence base. Clinicians are faced with multiple potential pharmacologic choices—clozapine, quetiapine, pimavanserin, and occasionally other antipsychotics—yet the relative efficacy and safety of these agents remain unclear [11]. Head-to-head RCTs directly comparing treatments are rare, and most trials have been placebo-controlled with modest sample sizes. Consequently, treatment decisions often rely on indirect comparisons, expert opinion, or local prescribing practices, leading to substantial variability in care. The absence of a comprehensive, evidence-based ranking of available therapies poses a barrier to guideline development and informed clinical decision-making.

This is where **network meta-analysis (NMA)** becomes particularly valuable. Unlike traditional pairwise meta-analysis, NMA allows simultaneous synthesis of direct and indirect evidence across a network of interventions. By integrating data from trials with different comparators, NMA provides estimates of relative efficacy and safety even in the absence of direct head-to-head studies. Moreover, NMA can generate a treatment hierarchy, ranking interventions according to their probability of being most effective or safest. Bayesian approaches to NMA further allow probabilistic interpretation of results, which may be more intuitive for clinicians.

The application of NMA in PD psychosis is especially timely. With the expanding range of pharmacologic options, including novel agents like pimavanserin, stakeholders require robust comparative evidence to inform practice and policy. Patients, caregivers, and clinicians must navigate complex trade-offs between psychosis control, preservation of motor function, and avoidance of serious adverse events. Health systems also need comparative data to guide formulary decisions and resource allocation. A rigorous synthesis of existing randomized trial evidence through NMA can fill these gaps by clarifying relative benefits and risks, identifying the most promising therapies, and highlighting areas where evidence remains insufficient.

In summary, psychosis is a highly prevalent and distressing complication of PD that substantially undermines quality of life and complicates disease management. Current treatment options are limited, variably effective, and associated with important safety concerns. Clozapine is effective but impractical for widespread use; quetiapine is widely prescribed but of uncertain efficacy; pimavanserin is promising but requires further comparative evaluation. Direct head-to-head trial evidence is scarce, leaving clinicians with limited guidance. By harnessing the power of network meta-analysis, this review seeks to provide a comprehensive, comparative assessment of pharmacologic therapies for Parkinson’s disease psychosis, with the ultimate goal of guiding evidence-based clinical practice and future research priorities.

## Methods

### 2.1 Literature Search

This meta-analysis was conducted according to a prespecified protocol and reported in accordance with PRISMA guidelines [12]. The Search was conducted using comprehensive databases like PubMed, Scopus, Embase, and CENTRAL till August 2025. The study protocol is registered with PROSPERO (International Prospective Register of Systematic Reviews) under registration number CRD420251143957. The search targeted studies meeting prespecified eligibility criteria—evaluations of the efficacy and safety of atypical anti-psychotics in psychosis during Parkinson’s. Guided by the PICOS framework, we used the Boolean expression (“Parkinson Disease”[Mesh] OR “Parkinson*”) AND (psychosis OR hallucinat* OR delusion*) AND (pimavanserin OR clozapine OR quetiapine OR olanzapine OR risperidone OR “antipsychotic*”) Reference lists of eligible and pertinent articles were hand-searched to identify additional records. No language limits were applied. Full search string is in Supplementary File.

### 2.2 Screening

Two reviewers independently screened all retrieved records in a two-stage process. First, titles and abstracts were assessed for relevance against the predefined eligibility criteria. Potentially eligible studies then underwent full-text review to confirm inclusion. Disagreements at either stage were resolved through discussion, and if necessary, by consultation with a third reviewer. Reasons for exclusion at the full-text stage were documented. The process was managed using reference management software to identify duplicates and ensure consistency. Screening decisions were recorded in detail, and the overall selection process was summarized using a PRISMA 2020 flow diagram.

### 2.3 Data Extraction and Statistical Analysis

Two reviewers independently extracted study characteristics, participant demographics, interventions, and outcomes using a standardized form. Continuous outcomes (e.g., BPRS, CGI, UPDRS) were recorded as means and SDs; binary outcomes (e.g., adverse events, discontinuations, mortality) as events and totals. A random-effects network meta-analysis was conducted, estimating mean differences or standardized mean differences for continuous data and risk ratios for binary outcomes. Multi-arm trials were adjusted for correlations, with placebo as the common comparator. Consistency was examined using design-by-treatment interaction and node-splitting models. Treatments were ranked by SUCRA and P-scores.

### 2.4 Risk of Bias Assessment

Two investigators independently assessed risk of bias using the Cochrane Risk of Bias 2 (RoB 2) tool for randomized trials [13]. Certainty of evidence for each prespecified outcome was appraised with the GRADE framework, considering risk of bias, inconsistency, indirectness, imprecision, and publication bias, and categorized as high, moderate, or low.

## Results

### Demographics

A total of 3632 studies were analyzed out of which, 1816 were removed because of duplicates, whereas, 672 were marked as 672 ineligible, and records 893 were removed for other reasons. The total records were screened 251. 109 records were excluded, thus 142 were sought for retrieval, out of which 68 were not retrieved. 74 studies were analysed for eligibility, out of which 26 were abstracts, 26 were cohort studies, leading to inclusion of 22 studies (Figure 1) [14–35]. A total of 2,047 patients with Parkinson’s disease psychosis were included across randomized controlled trials. The mean age of participants was 70.6 years (±12), reflecting an elderly population typically affected by late-stage disease. The overall cohort comprised 1,038 males and 997 females, indicating a nearly balanced sex distribution. The mean follow-up duration was 2.73 months (±1), consistent with short-to medium-term trial designs. Across treatment arms, 1,091 patients received active interventions and 1,036 patients were assigned to control groups. Intervention subgroups included clozapine (n=139), olanzapine (n=111), pimavanserin (n=746), quetiapine (n=126), risperidone (n=5), and ziprasidone (n=8), compared with placebo (n=992). Table S1.

**Figure 1.**
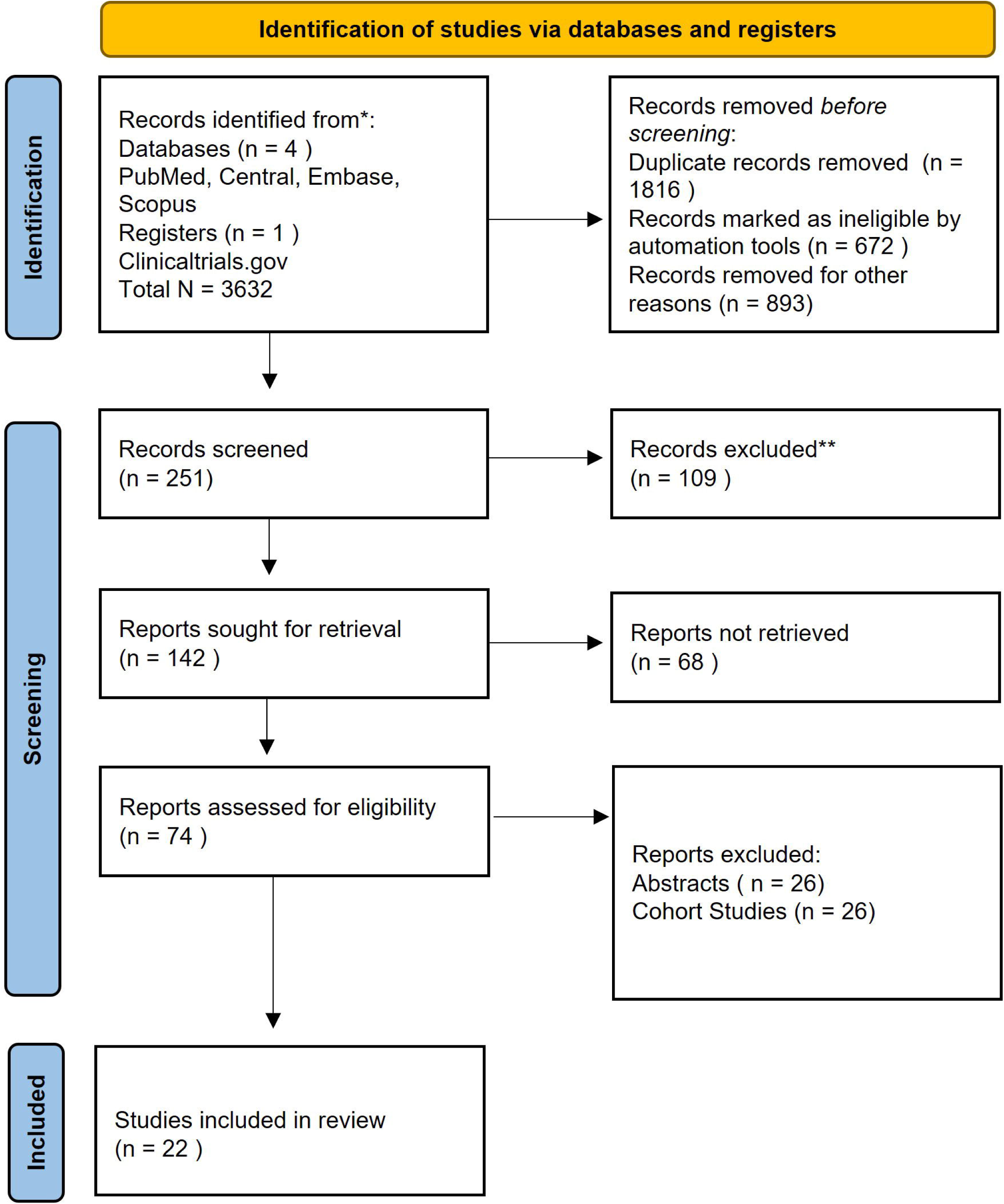
PRISMA Flow Diagram

### BPRS Outcomes of each treatment

The forest plot comparing treatments with placebo for changes in Brief Psychiatric Rating Scale (BPRS) scores showed that none of the antipsychotics demonstrated statistically significant superiority. Clozapine indicated the greatest potential improvement (mean difference –10.0, 95% CrI –27.2 to 7.27), though the interval overlapped zero, reflecting uncertainty. Quetiapine (–7.12, 95% CrI –20.0 to 5.43) and risperidone (–15.8, 95% CrI –48.9 to 16.9) also suggested possible benefit, but wide intervals indicated imprecision, particularly for risperidone due to small sample size. Olanzapine showed minimal effect (–0.686, 95% CrI – 17.5 to 16.2). Overall, evidence remains inconclusive regarding BPRS improvement with these agents. The SUCRA ranking plot shows risperidone and clozapine as the most effective treatments for BPRS improvement, while olanzapine and placebo ranked lowest. Table S2, Figure 2, Figure 3, Figure S1.

**Figure 2.**
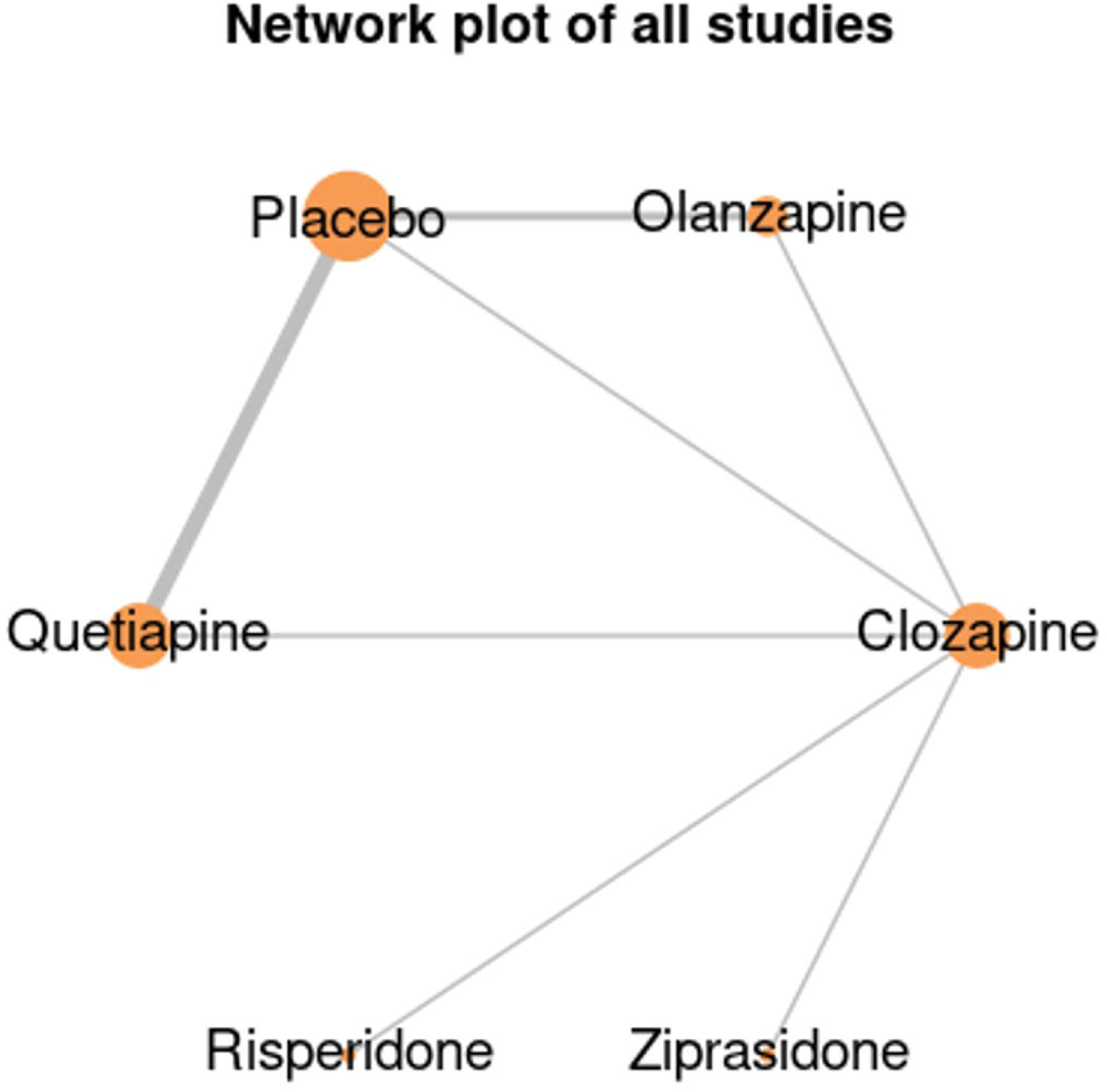
Network Diagram of all treatments for BPRS.

**Figure 3.**
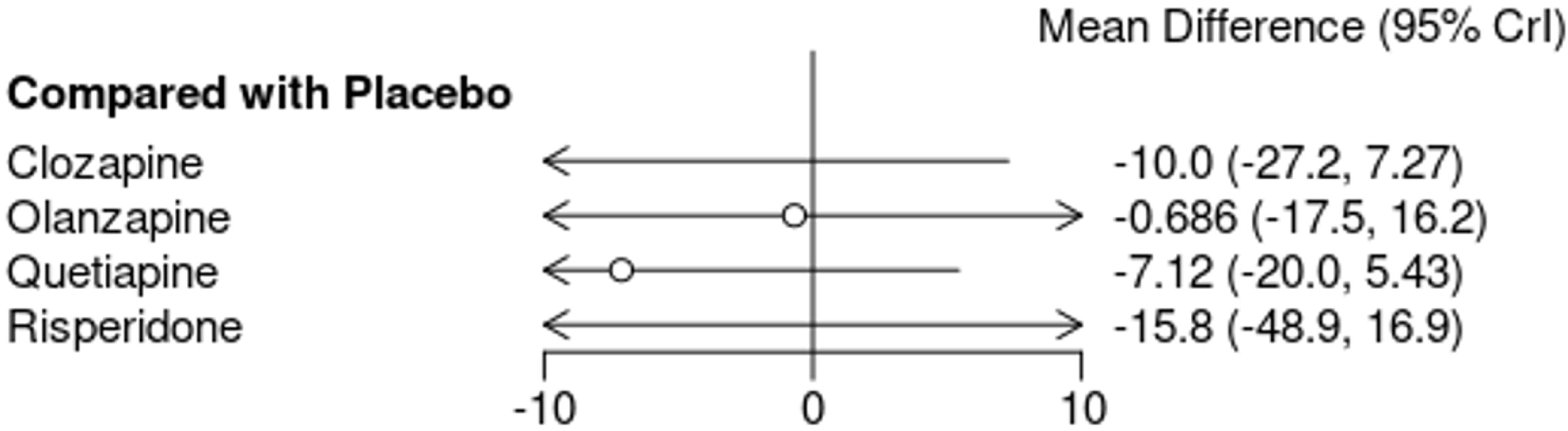
Forest Plot of all treatments for BPRS (Mean Difference and SD).

The forest plot summarizes the effects of clozapine, olanzapine, and quetiapine compared with placebo on Brief Psychiatric Rating Scale (BPRS) scores. Clozapine showed mixed findings across trials, with effect estimates ranging from improvement to worsening, resulting in an overall non-significant effect (mean difference –3.58, 95% CI –21.90 to 14.75). Olanzapine demonstrated inconsistent results, with pooled analysis indicating no clear benefit (2.43, 95% CI –2.52 to 7.38). Quetiapine trials similarly varied, producing a pooled effect that did not reach statistical significance (–5.25, 95% CI –16.62 to 6.13). The overall analysis suggested no significant reduction in BPRS scores across treatments (–2.51, 95% CI –8.95 to 3.94), with high heterogeneity observed among studies. Figure 4.

**Figure 4.**
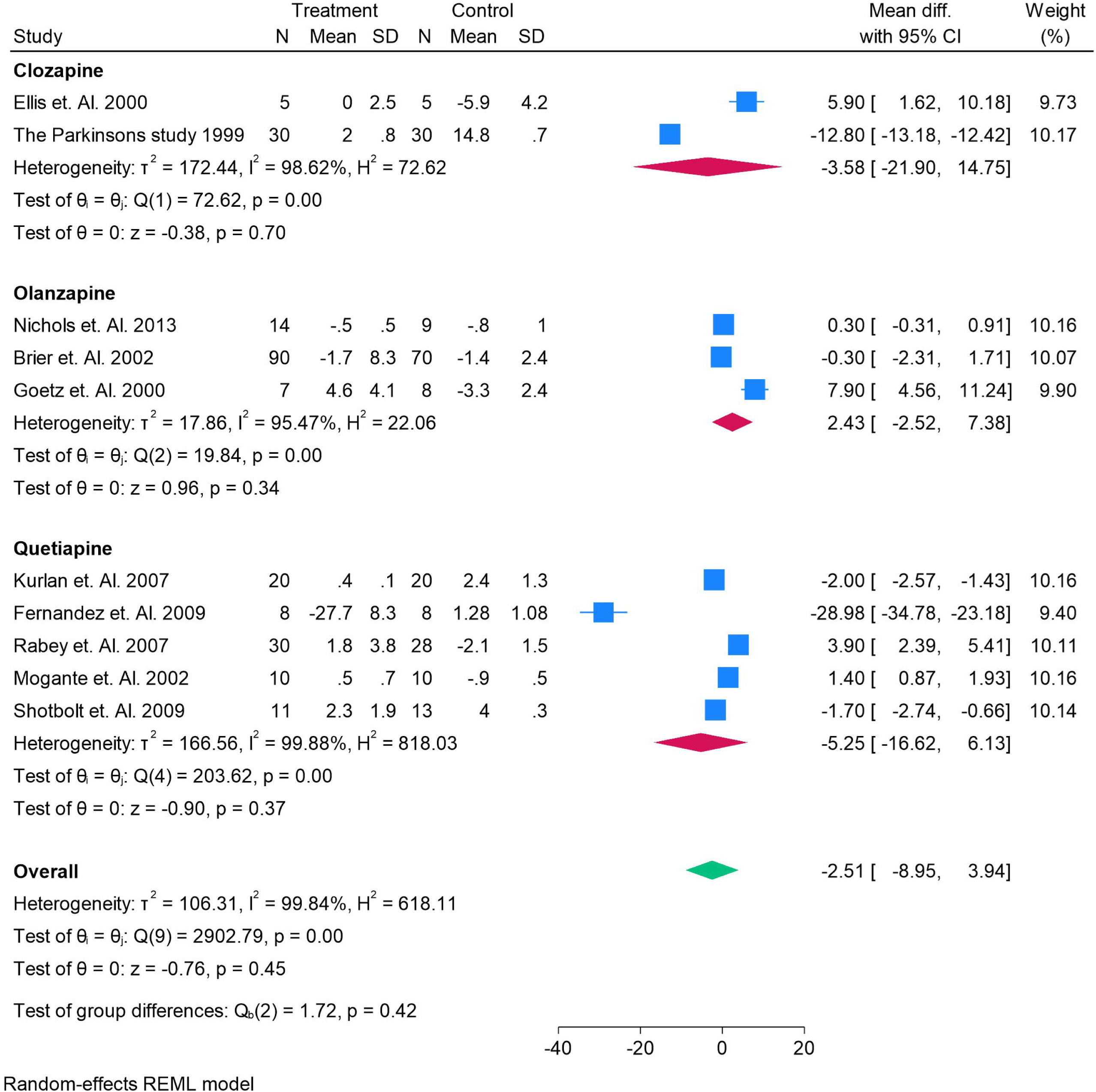
Forest Plot of all treatments for BPRS (Mean Difference and SD).

### CGI-S Outcomes of each treatment

The forest plot displays the comparative effects of antipsychotics versus placebo on the Clinical Global Impression–Severity (CGI-S) scale. None of the agents showed statistically significant superiority, as all 95% credible intervals crossed zero. Clozapine (–1.21, 95% CrI –3.81 to 1.39), olanzapine (–1.90, 95% CrI –4.49 to 0.72), quetiapine (–1.08, 95% CrI –3.80 to 1.66), risperidone (–2.01, 95% CrI –15.1 to 10.5), and ziprasidone (–1.41, 95% CrI –5.44 to 2.60) suggested possible reductions in severity, though imprecise.

Pimavanserin showed a slight nonsignificant increase (0.429, 95% CrI –1.09 to 1.94). Overall, evidence indicates no clear treatment benefit over placebo. The SUCRA ranking plot for CGI-S outcomes indicates that **olanzapine** had the highest probability of being the most effective treatment, followed by **ziprasidone, clozapine, risperidone, and quetiapine**, which showed moderate rankings. **Placebo** ranked lower, while **pimavanserin** had the lowest SUCRA score, suggesting the least likelihood of benefit in reducing CGI-S severity. Figure 5, Figure 6, Figure S2, Table S2.

**Figure 5.**
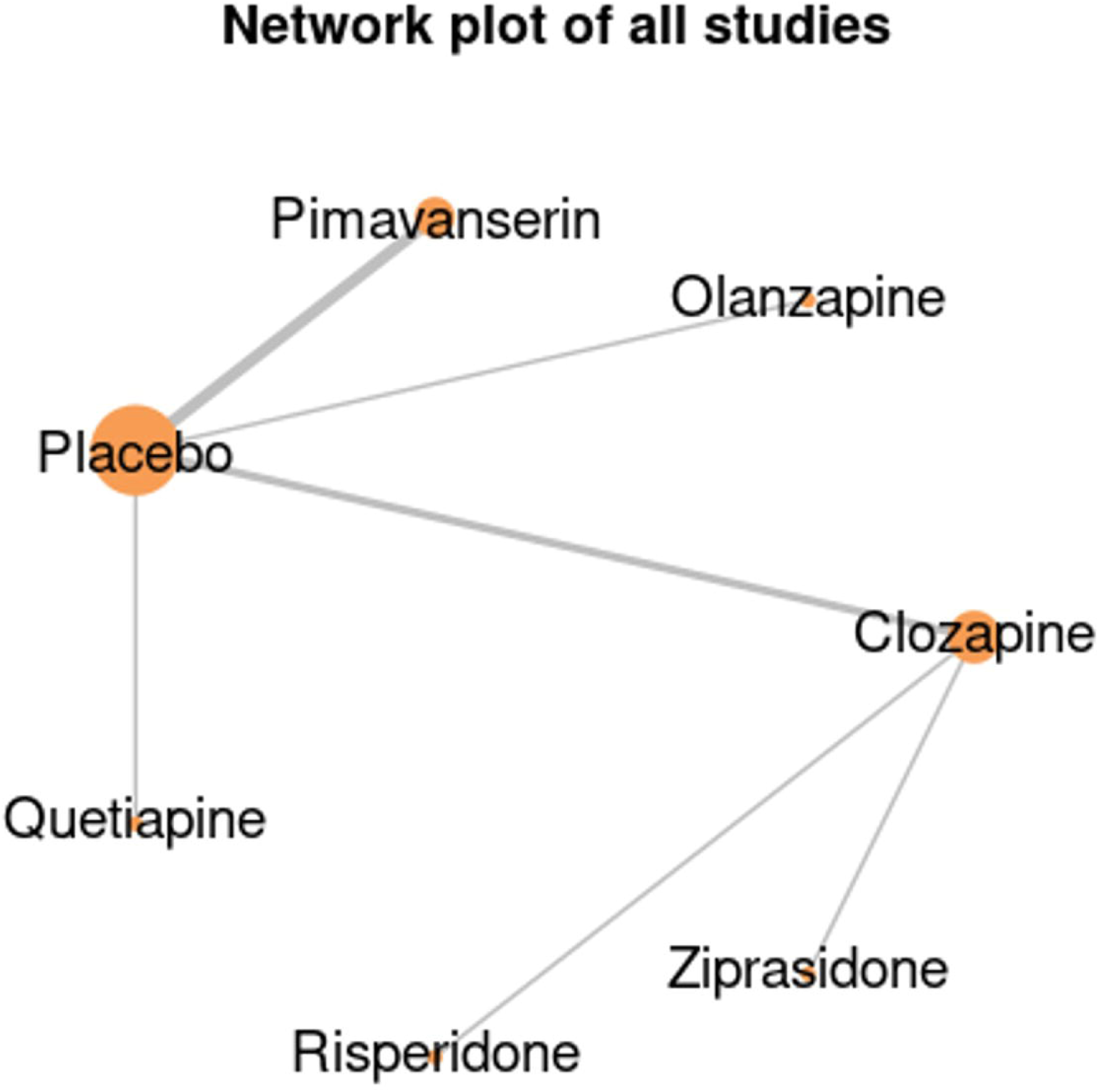
Network Diagram of all treatments for CGI-S.

**Figure 6.**
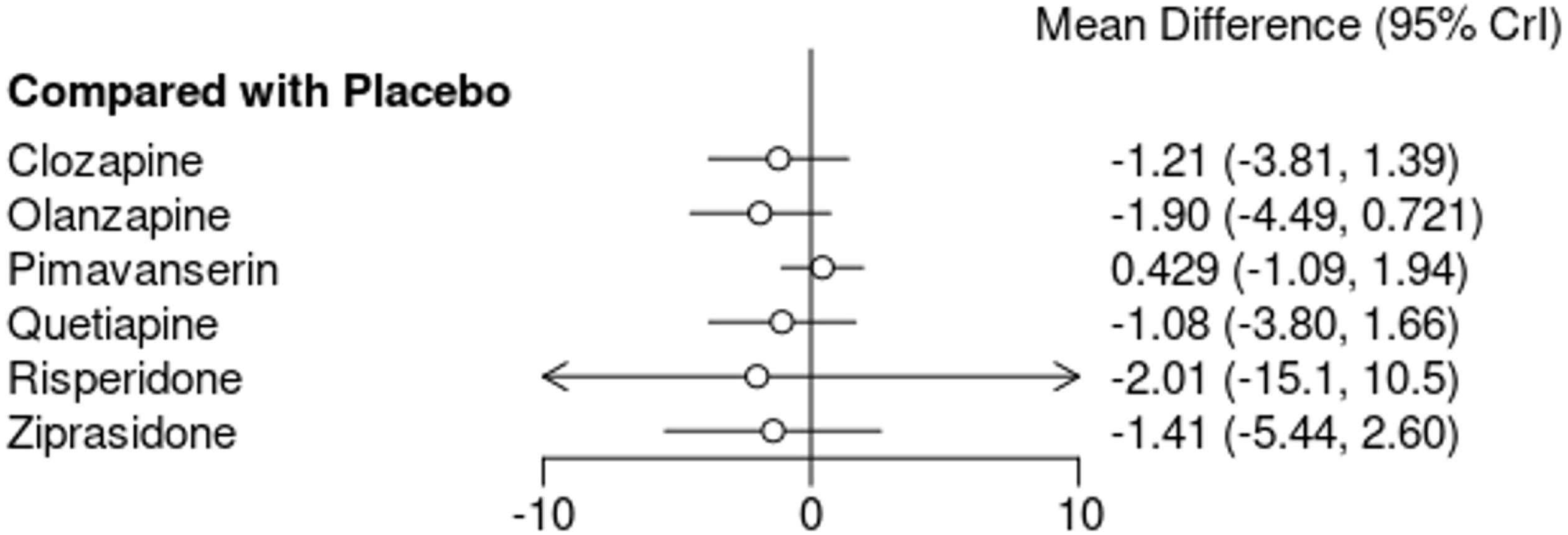
Forest Plot of all treatments for CGI-S (Mean Difference and SD).

The forest plot presents the effects of clozapine, olanzapine, pimavanserin, quetiapine, and ziprasidone on CGI-S scores compared with placebo. Clozapine demonstrated a consistent reduction in severity with a pooled mean difference of –1.10 (95% CI –1.30 to –0.91). Olanzapine showed variable effects across studies, with no significant overall benefit (–1.10, 95% CI –2.67 to 0.47). Pimavanserin yielded mixed findings, with newer trials showing slight improvement, though the pooled effect was non-significant (0.43, 95% CI –0.80 to 1.66). Quetiapine demonstrated a significant reduction (–1.09, 95% CI –1.97 to –0.21), while ziprasidone showed no effect (–0.20, 95% CI –1.96 to 1.56). Overall, pooled analysis indicated no clear superiority of treatments (–0.49, 95% CI –1.19 to 0.20), with substantial heterogeneity. Figure 7.

**Figure 7.**
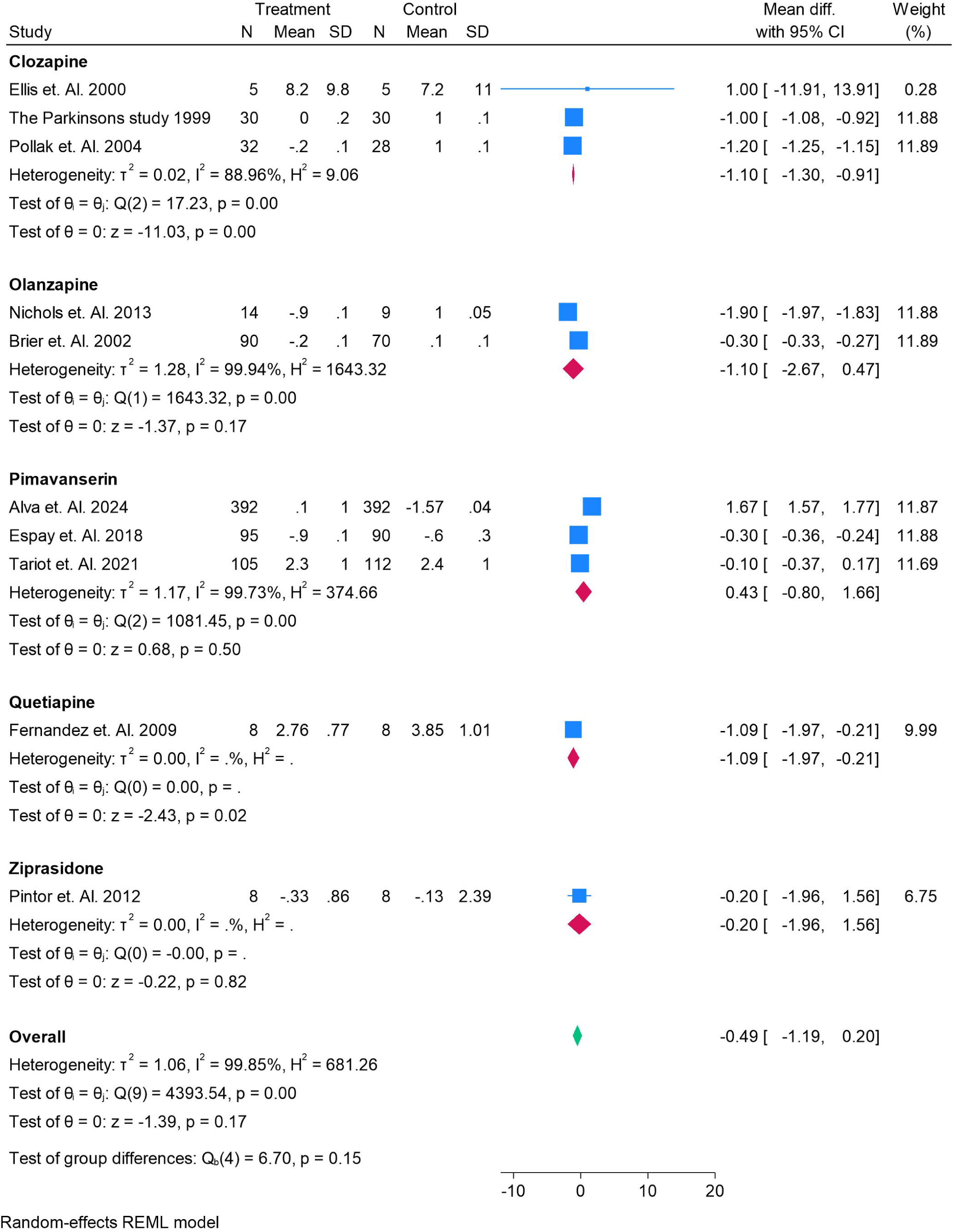
Forest Plot of all treatments for CGI-S(Mean Difference and SD).

### UPDRS-II Outcomes of each treatment

The forest plot illustrates the effects of antipsychotic treatments compared with placebo on UPDRS-II (activities of daily living) scores. None of the drugs demonstrated a statistically significant impact, as all 95% credible intervals crossed zero. Clozapine (–0.82, 95% CrI –4.57 to 2.96), olanzapine (–0.31, 95% CrI –6.76 to 5.14), quetiapine (–0.52, 95% CrI –3.94 to 2.50), and ziprasidone (–3.88, 95% CrI –17.7 to 10.1) suggested possible motor worsening, though imprecisely estimated. Pimavanserin (0.16, 95% CrI –5.17 to 5.52) showed no meaningful difference. Risperidone indicated a positive change (3.37, 95% CrI –6.65 to 13.5), but with wide uncertainty. Overall, no treatment significantly altered UPDRS-II outcomes. The SUCRA ranking plot for UPDRS-II outcomes shows that **ziprasidone** had the highest probability of being the most favorable treatment, followed closely by **clozapine, quetiapine, and olanzapine**. **Placebo** and **pimavanserin** occupied intermediate ranks, while **risperidone** had the lowest SUCRA score, indicating the least likelihood of benefit for activities of daily living in patients with Parkinson’s disease psychosis. Figure 7, 8, 9, Figure S3, Table S3.

The forest plot summarizes the effects of clozapine, olanzapine, pimavanserin, quetiapine, and ziprasidone on UPDRS-II (activities of daily living) scores compared with placebo. Clozapine showed mixed results across trials, with no significant overall effect (–0.41, 95% CI –2.97 to 2.16). Olanzapine also demonstrated variable findings, yielding a non-significant pooled mean difference (–1.46, 95% CI –10.85 to 7.94). Pimavanserin suggested no meaningful change (0.34, 95% CI –0.51 to 1.19). Quetiapine trials showed heterogeneous effects, with the pooled estimate again non-significant (0.35, 95% CI –2.22 to 2.91). Ziprasidone demonstrated a potential negative effect, though with wide uncertainty (–2.90, 95% CI –14.21 to 8.41). Overall, pooled analysis indicated no significant differences between active treatments and placebo (–0.02, 95% CI –1.43 to 1.40), with substantial heterogeneity across studies. Figure 10.

**Figure 8.**
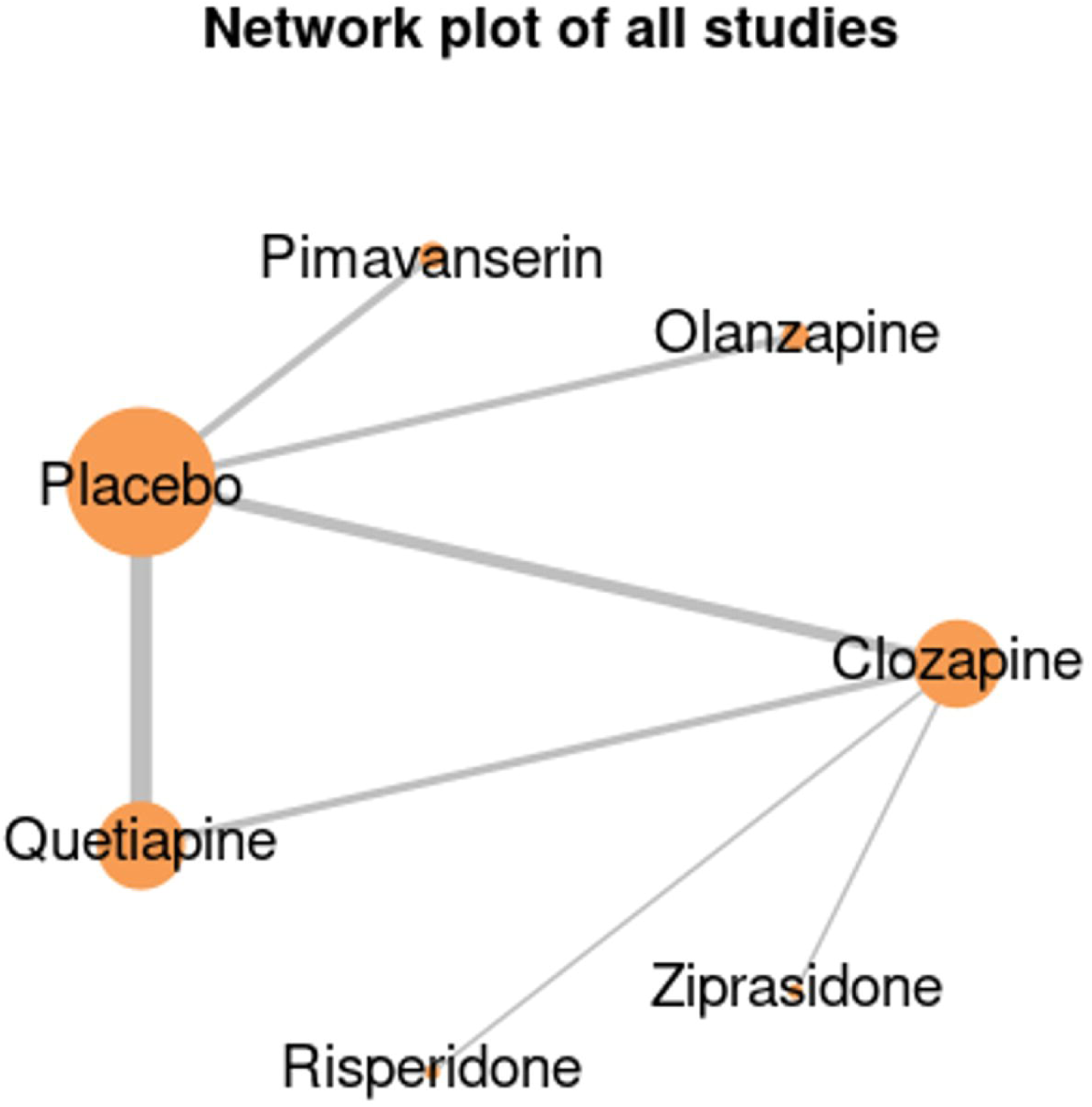
Network Diagram of all treatments for UPDRS-11

**Figure 9.**
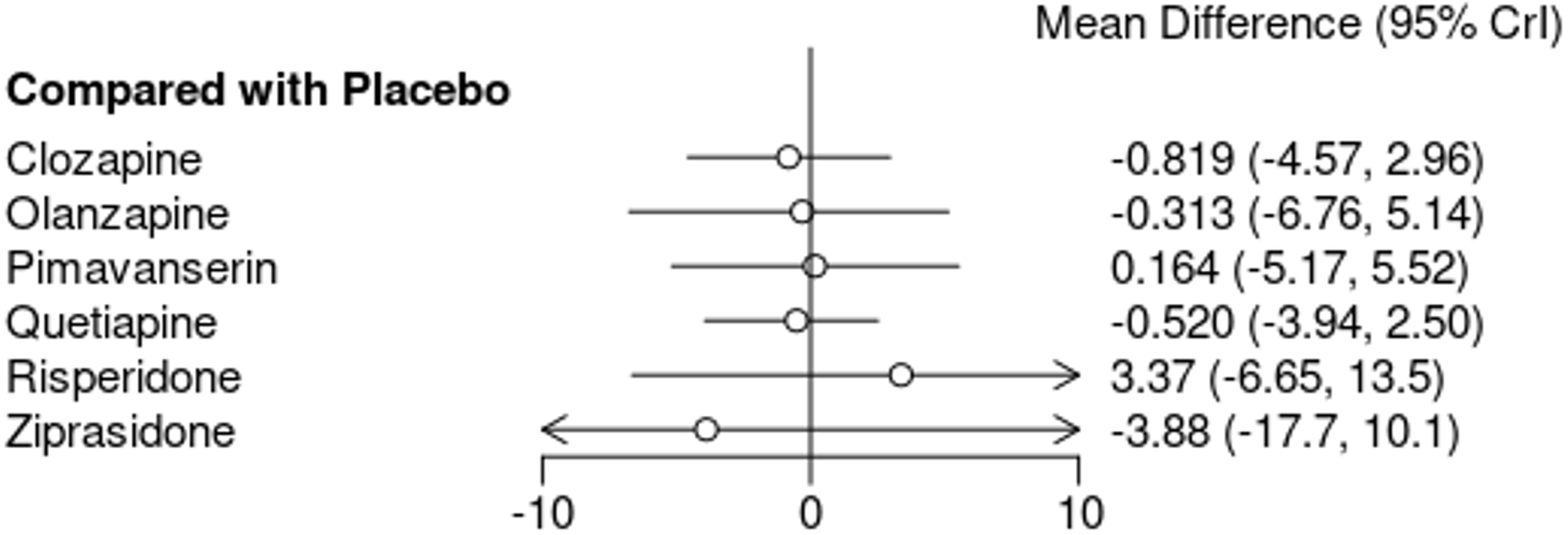
Forest Plot of all treatments for UPDRS-II (Mean Difference and SD).

**Figure 10.**
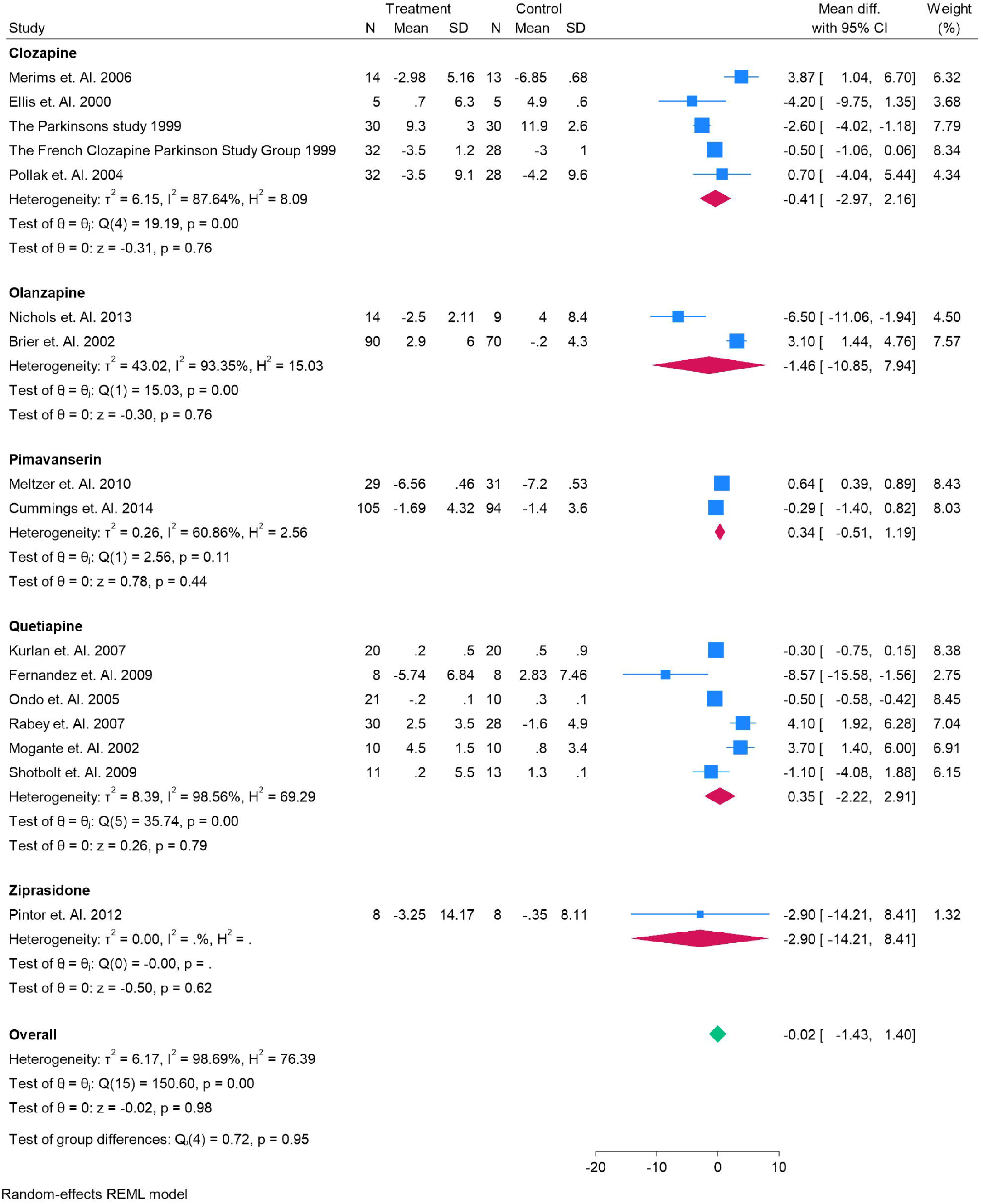
Forest Plot of all treatments for UPDRS-11 (Mean Difference and SD).

### Adverse Events

#### Worsening Parkinsons Disease

The forest plot presents the risk of worsening Parkinson’s disease (PD) symptoms as an adverse event across antipsychotic treatments compared with placebo. Clozapine showed a pooled risk ratio of 0.51 (95% CI – 0.16 to 1.18), suggesting no significant association. Olanzapine (0.35, 95% CI –0.78 to 1.48), pimavanserin (–0.08, 95% CI –0.54 to 0.37), quetiapine (0.04, 95% CI –0.79 to 0.87), and ziprasidone (–0.69, 95% CI – 2.08 to 0.69) all demonstrated non-significant effects with confidence intervals crossing zero. The overall pooled analysis (0.05, 95% CI –0.24 to 0.35) indicated no meaningful increase in PD worsening with any treatment, with low heterogeneity. Figure 11.

**Figure 11.**
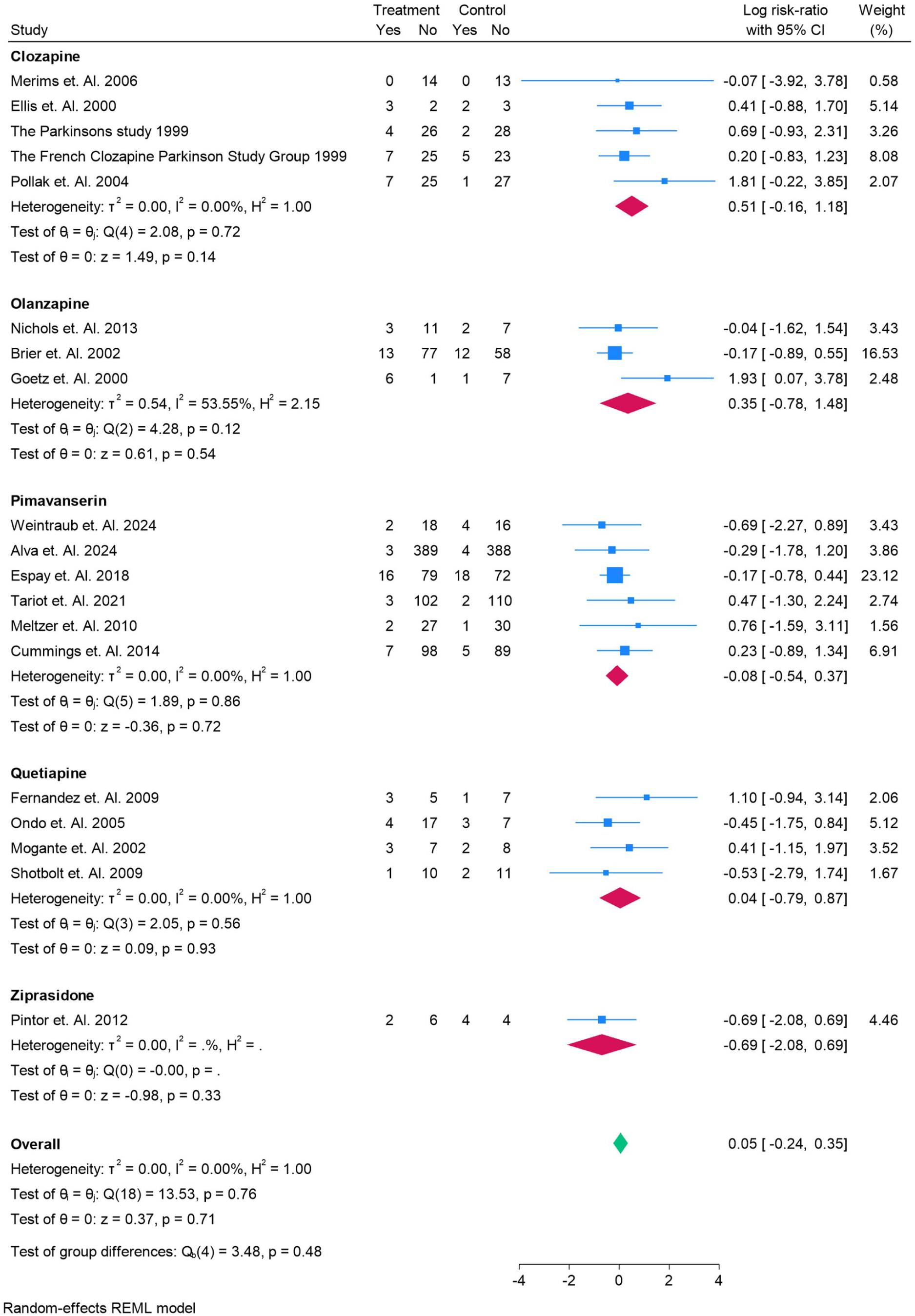
Worsening PD for Treatments as Risk Ratios for Adverse Events.

#### Insomnia

The forest plot shows the risk of insomnia as an adverse event across antipsychotic treatments compared with placebo. Clozapine demonstrated no significant association with insomnia (log risk ratio –0.00, 95% CI –0.49 to 0.49). Olanzapine also showed no significant effect (–0.17, 95% CI –1.09 to 0.76). Pimavanserin was associated with a slight but non-significant increase in risk (0.21, 95% CI –0.46 to 0.89). Quetiapine (0.02, 95% CI –0.85 to 0.90) and ziprasidone (–0.69, 95% CI –2.08 to 0.69) both showed no significant impact. Overall, pooled analysis (–0.01, 95% CI –0.33 to 0.32) indicated no meaningful increase in insomnia risk, with no heterogeneity across studies. Figure 12.

**Figure 12.**
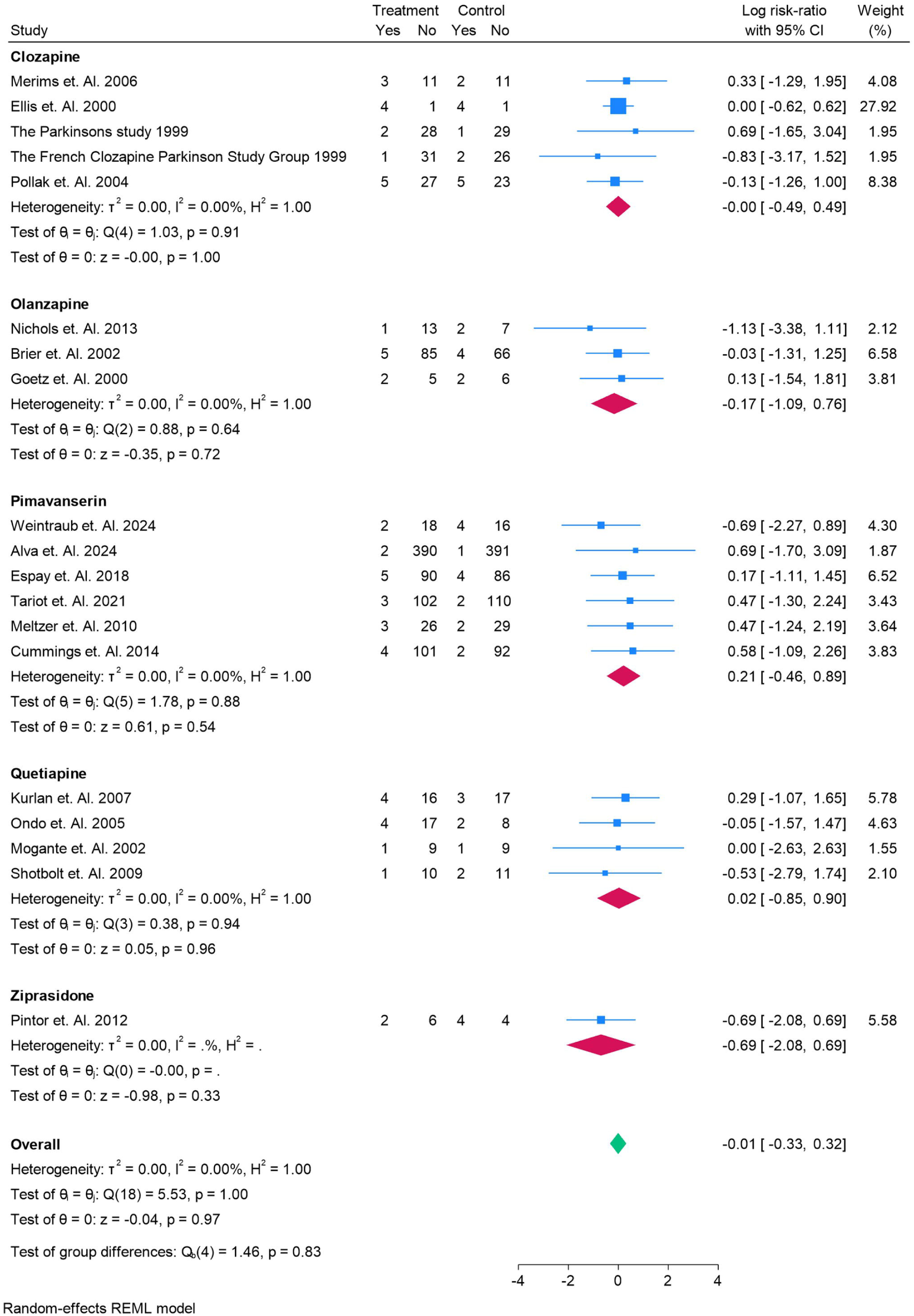
Insomnia for Treatments as Risk Ratios for Adverse Events.

#### Cardiovascular Events

The forest plot evaluates the risk of cardiovascular events across antipsychotic treatments compared with placebo. Clozapine showed no significant effect, with a pooled log risk ratio of –0.01 (95% CI –0.08 to 0.06). Olanzapine similarly demonstrated no association (0.02, 95% CI –0.05 to 0.09). Pimavanserin yielded a near-null effect (–0.00, 95% CI –0.01 to 0.01), while quetiapine showed a slight but non-significant increase (0.01, 95% CI –0.06 to 0.08). Ziprasidone data were limited, with no observed difference (0.00, 95% CI –0.57 to 0.57). The overall pooled estimate (–0.00, 95% CI –0.01 to 0.01) indicated no meaningful increase in cardiovascular risk, with no evidence of heterogeneity across studies. Figure 13.

**Figure 13.**
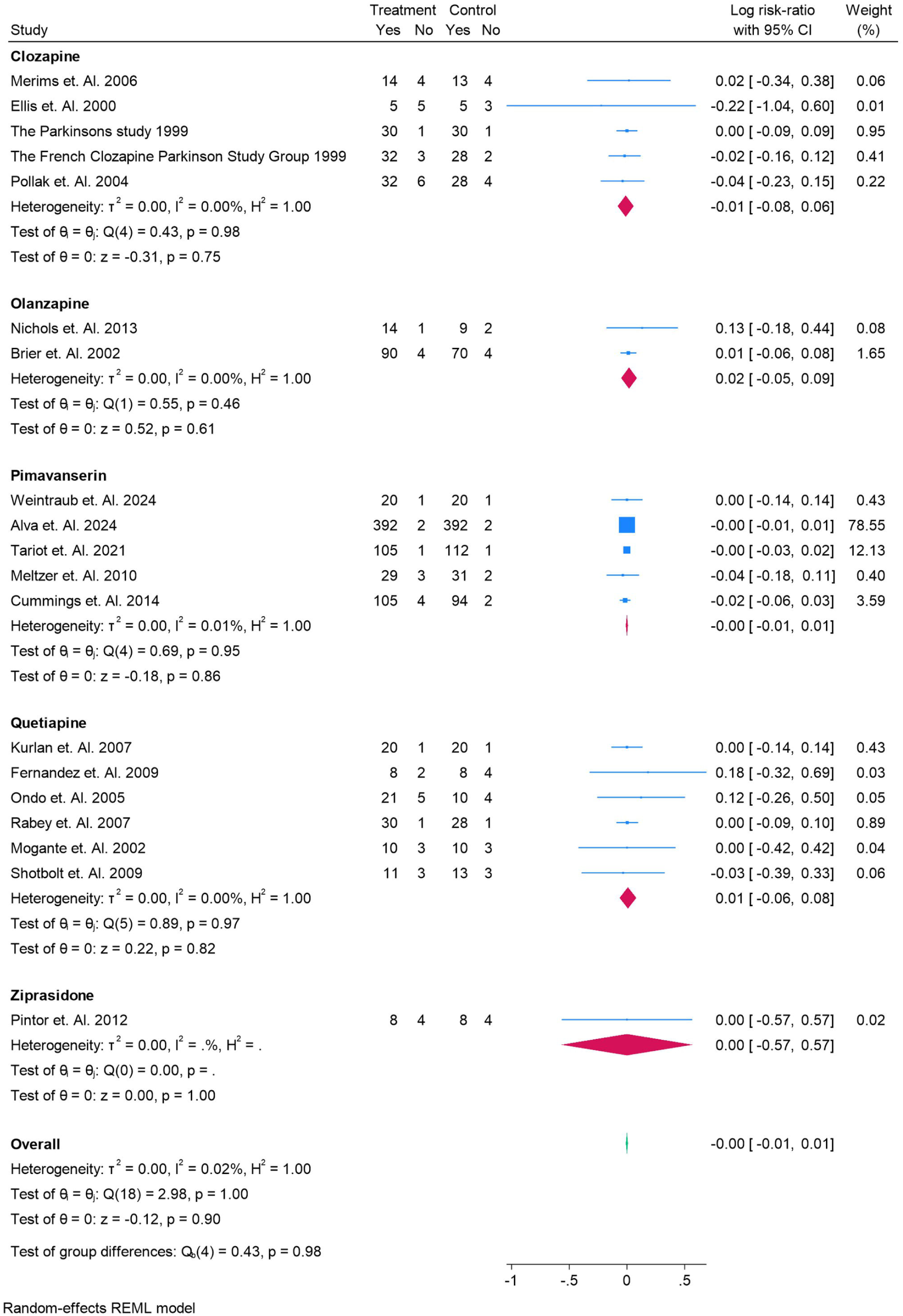
Cardiovascular Events for Treatments as Risk Ratios for Adverse Events.

#### Mortality

The forest plot evaluates mortality risk across antipsychotic treatments compared with placebo. Clozapine showed no association with mortality (log risk ratio –0.00, 95% CI –0.06 to 0.05), and olanzapine similarly demonstrated no significant effect (0.04, 95% CI –0.21 to 0.28). Pimavanserin produced consistently null findings across multiple trials, with an overall pooled estimate of –0.00 (95% CI –0.01 to 0.01). Quetiapine also revealed no significant impact (0.01, 95% CI –0.08 to 0.10). The overall analysis (–0.00, 95% CI –0.01 to 0.01) indicated no meaningful increase in mortality risk with any treatment, with zero heterogeneity across studies. Figure 14.

**Figure 14.**
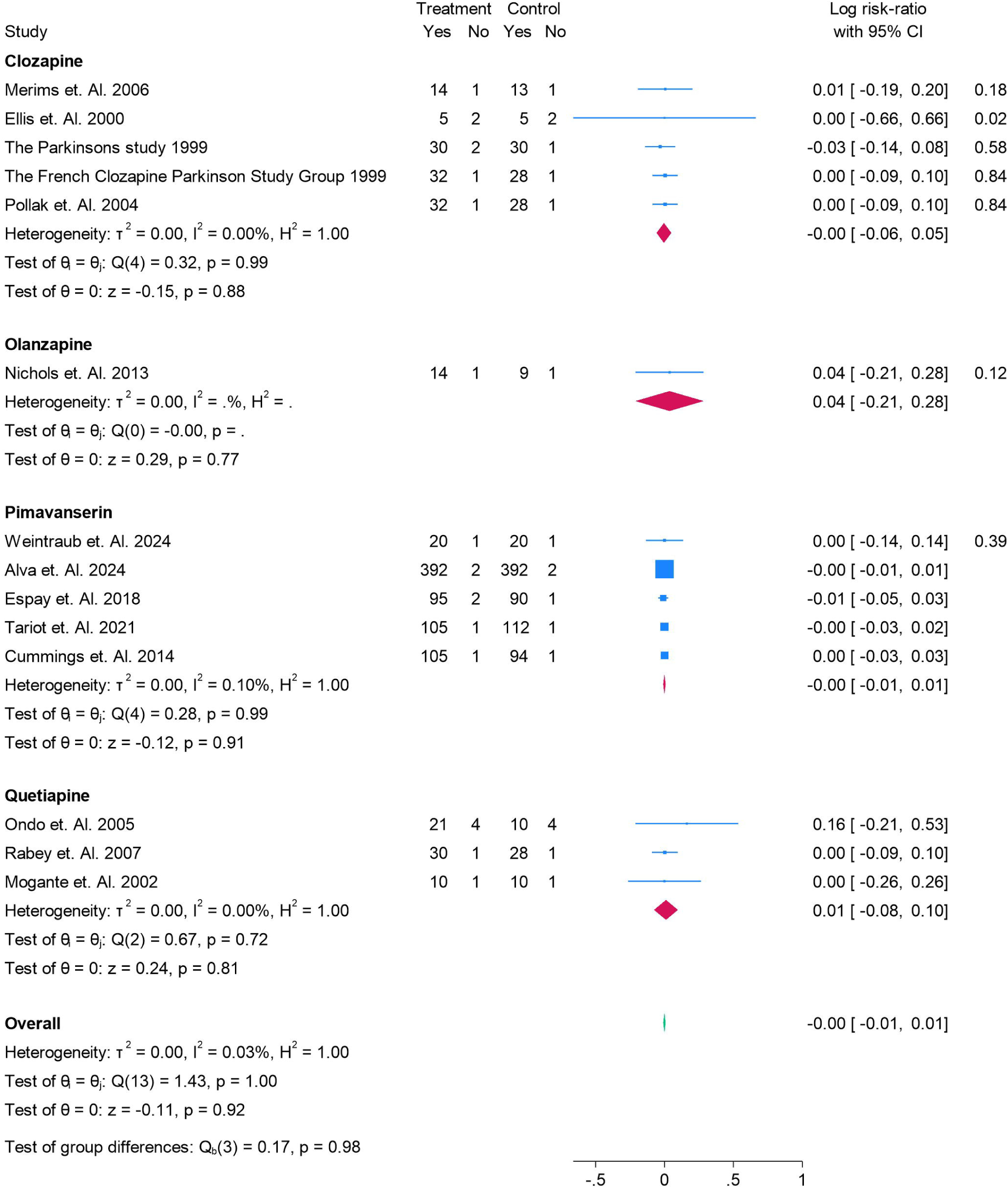
Mortality Events for Treatments as Risk Ratios for Adverse Events.

## Discussion

This network meta-analysis comprehensively evaluated the efficacy and safety of pharmacologic interventions for Parkinson’s disease psychosis (PDP) across randomized controlled trials. By incorporating both direct and indirect evidence, we were able to generate comparative estimates between multiple agents, including clozapine, quetiapine, olanzapine, risperidone, ziprasidone, and pimavanserin. Overall, our analysis suggests that no antipsychotic demonstrated consistent and statistically significant superiority over placebo across psychosis rating scales (BPRS, CGI-S). Clozapine and quetiapine showed trends toward psychotic symptom improvement, while risperidone and olanzapine offered no clear benefits and carried the risk of motor worsening. Pimavanserin, despite being the only FDA-approved treatment, showed modest effects on global severity scales but was not consistently superior to placebo. Importantly, none of the agents demonstrated an increased risk of mortality or cardiovascular events, although heterogeneity and wide confidence intervals underscore persistent uncertainty.

### Comparison with Previous Meta-analyses

Our findings align with earlier pairwise meta-analyses. A study concluded that clozapine was effective but limited by hematologic monitoring requirements, while quetiapine lacked convincing efficacy despite widespread use [36]. Similarly, a Cochrane review found strong evidence for clozapine but insufficient support for quetiapine, with olanzapine associated with motor deterioration [37]. A more recent systematic review echoed these concerns, highlighting pimavanserin as a promising option but stressing the need for long-term safety data [38].

Unlike these traditional meta-analyses, our NMA enables a unified ranking of agents. Clozapine and risperidone ranked highest for psychosis improvement on BPRS and CGI-S, although the latter’s profile is clouded by motor side effects in practice. Pimavanserin consistently ranked mid-tier, reflecting modest efficacy and tolerability advantages but not a clear superiority signal. Quetiapine and olanzapine clustered in lower ranks, consistent with their inconclusive or unfavorable efficacy data. Notably, our results extend the literature by incorporating ziprasidone, albeit with limited trial data.

### Clinical Implications

The findings reinforce clozapine as the most efficacious option, yet its hematologic monitoring requirements restrict routine use. Pimavanserin offers a mechanistically distinct alternative without dopaminergic antagonism, but its clinical effect sizes appear smaller than initially anticipated. Quetiapine remains widely prescribed, likely due to ease of use and safety perception, but our analysis confirms its lack of robust efficacy. Olanzapine and risperidone should be avoided given risks of motor worsening, despite their relatively favorable ranking in indirect analyses. From a safety standpoint, none of the agents demonstrated excess mortality or cardiovascular risk, though the short follow-up durations (mean 2.7 months) limit conclusions about long-term harm.

### Strengths and Limitations

The strengths of this review include the comprehensive search strategy, application of network meta-analysis to integrate direct and indirect comparisons, and systematic assessment of efficacy, safety, and tolerability outcomes. By aligning multiple psychosis and motor function scales, we minimized bias from outcome heterogeneity and provided clinically meaningful rankings.

However, several limitations must be acknowledged. The number of included trials was small for several agents, especially risperidone and ziprasidone, leading to imprecision in estimates. Follow-up durations were short, restricting assessment of sustained efficacy and long-term safety. Significant heterogeneity was observed in some analyses, particularly for BPRS outcomes, reflecting variability in trial populations, outcome measures, and dosing regimens. Furthermore, indirect comparisons rely on the assumption of transitivity, which may be challenged by differences in baseline severity, concomitant dopaminergic therapy, and trial design.

### Future Directions

Further large-scale, head-to-head RCTs with longer follow-up are needed to clarify the comparative effectiveness of pimavanserin against clozapine and quetiapine, the two most clinically relevant comparators. Trials should also systematically evaluate patient-centered outcomes such as quality of life, caregiver burden, and functional capacity. Real-world evidence from registries and claims data will be important to contextualize trial findings and assess safety signals over extended periods.

## Conclusion

In conclusion, this network meta-analysis highlights the persistent therapeutic challenges in treating PDP. Clozapine remains the most effective antipsychotic but is underutilized due to safety monitoring requirements. Pimavanserin offers an alternative with an improved safety profile, though its efficacy appears modest. Quetiapine, despite frequent use, is not supported by strong evidence, and olanzapine and risperidone are limited by motor worsening risks. Our findings emphasize the need for more robust comparative trials to guide evidence-based treatment strategies for PDP.

## Authors’ contributions

All authors contributed equally to the manuscript and read and approved the final version of the manuscript.

## Data Availability

Supplementary file

