## supplementary file for "Comparative Efficacy and Safety of Antipsychotics for Parkinson’s Disease Psychosis: A Systematic Review and Network Meta-Analysis"

Supplementary Files

Search String

("Parkinson Disease"[Mesh] OR parkinson* OR "PD") AND (psychosis OR hallucinat* OR delusion* OR "SAPS-PD" OR "BPRS" OR "CGI") AND (pimavanserin OR clozapine OR quetiapine OR olanzapine OR risperidone OR ziprasidone OR aripiprazole OR "typical antipsychotic*" OR "antipsychotic*")

Table S1. Demographics and Summary Findings

| **Author Name year** | **Country of Trial** | **Total Sample** | **Male** | **Female** | **Mean Age** | **Follow Up Time** | Psychosis definition | **Rx Name** | **Rx Sample** | **Cx Name** | **Cx Sample** | **GRADE** | Main Finding |
| --- | --- | --- | --- | --- | --- | --- | --- | --- | --- | --- | --- | --- | --- |
| 14 Merims et. Al. 2006 | Israel | 27 | 16 | 11 | 72.8 | 5.5 | Recent-onset, clinically significant psychotic symptoms (hallucinations and/or delusions) in PD requiring neuroleptic treatment, assessed with NPI hallucination and delusion items. | Clozapine | 14 | Quetiapine | 13 | High | Both clozapine and quetiapine improved PD psychosis without worsening motor symptoms, with **clozapine showing greater reduction in delusion frequency (and a trend for hallucinations)** but at the cost of leukopenia risk. |
| 15 Kurlan et. Al. 2007 | USA | 40 | 25 | 15 | 73.5 | 2.5 | Presence of psychosis as defined by Jeste & Finkel (2000) and DSM-IV (or agitation per Cohen-Mansfield & Billig), in patients with dementia and parkinsonism. | Quetiapine | 20 | Placebo | 20 | High | Quetiapine was generally well-tolerated without worsening parkinsonism but did not show demonstrable efficacy over placebo for agitation/psychosis at the doses used. |
| 16 Weintraub et. Al. 2024 | Multicenter | 36 | 22 | 14 | 72.6 | 6 | Psychosis defined as **hallucinations and/or delusions**, assessed with **SAPS-H+D** (Hallucinations + Delusions subscales). | Pimavanserin | 20 | Placebo | 20 | High | In PDD with psychosis who first responded to open-label pimavanserin, **continuing pimavanserin 34 mg cut the risk of psychosis relapse by ~95% vs placebo** during randomized withdrawal (HR = 0.052; 95% CI 0.016–0.166). |
| 17 Fernandez et. Al. 2009 | USA | 16 | 8 | 8 | 68 | 3 | Consistent and persistent (>1 month), predominantly nocturnal visual hallucinations in PD. | Quetiapine | 8 | Placebo | 8 | High | Quetiapine improved visual hallucinations versus placebo (better CGI improvement and BPRS hallucination item), without significant change in REM sleep architecture or UPDRS motor worsening. |
| 18 Cortese et. Al. 2008 | Canada | 22 | 12 | 10 | 40.8 | 3 | diagnosis was DSM-IV schizophrenia and symptom severity was assessed with PANSS. | Quetiapine | 13 | Placebo | 9 | High | Switching to quetiapine significantly reduced extrapyramidal symptoms (parkinsonism and akathisia clinically; dyskinesia instrumentally) versus staying on prior antipsychotic over 3 months. |
| 19 Nichols et. Al. 2013 | USA | 24 | 12 | 12 | 72.6 | 1 | Clinically significant **hallucinations or delusions** in idiopathic PD on dopaminomimetics, as judged by clinicians/investigator. | Olanzapine | 14 | Placebo | 9 | High | Olanzapine (2.5–5 mg) **was not more effective than placebo** for PD psychosis and was associated with **more mild side-effects** (often motor worsening). |
| 20 Alva et. Al. 2024 | Multicenter | 784 | 331 | 453 | 72.4 | 4 | Patients required a neuropsychiatric symptom warranting antipsychotic treatment, evidenced by **NPI (frequency×severity) ≥4** in a domain such as **delusions or hallucinations**. | Pimavanserin | 392 | Placebo | 392 | High | Pimavanserin was **well tolerated** with **TEAEs similar to placebo** and **no motor or cognitive impairment**, supporting a favorable safety profile in elderly patients with NDD (including PD). |
| 21 Ellis et. Al. 2000 | USA | 10 | 5 | 5 | 74 | 3 | Levodopa-exacerbated psychosis not satisfactorily managed by levodopa dose reduction. | Clozapine | 5 | Risperidone | 5 | High | Clozapine and risperidone showed similar efficacy for PD psychosis on BPRS outcomes, but motor function tended to worsen more with risperidone, warranting caution. |
| 22 Ondo et. Al. 2005 | USA | 31 | 17 | 14 | 73 | 3 | PD subjects with **subjectively problematic visual hallucinations while on dopaminergic medications**. | Quetiapine | 21 | Placebo | 10 | High | Quetiapine up to 200 mg/day was **well tolerated and did not worsen UPDRS**, but **did not significantly improve psychosis ratings vs placebo** over 12 weeks. |
| 23 Espay et. Al. 2018 | USA | 185 | 116 | 69 | 72.4 | 1.5 | Hallucinations and/or delusions in PD, present ≥1 month and severe enough to require antipsychotic treatment (per NINDS/NIMH criteria). | Pimavanserin | 95 | Placebo | 90 | High | Pimavanserin significantly improved psychosis (SAPS-PD) versus placebo across cognition strata, with numerically larger effects in cognitively impaired patients and similar overall tolerability. |
| 24 The Parkinsons study 1999 | USA | 60 | 34 | 26 | 71.9 | 1 | A major psychiatric illness in which reality testing was impaired, typically by the presence of hallucinations or delusions, leading to substantial communication and social problems | Clozapine | 30 | Placebo | 30 | High | Low-dose clozapine (≤ 50 mg/day) significantly improved psychosis in PD without worsening parkinsonism and even reduced tremor. |
| 25 Tariot et. Al. 2021 | Multicenter | 217 | 86 | 131 | 73.8 | 4.1 | Dementia-related psychosis with hallucinations and/or delusions, requiring SAPS-H+D ≥10, SAPS-H+D global ≥4, CGI-S ≥4, and symptoms for ≥2 months at screening/baseline. | Pimavanserin | 105 | Placebo | 112 | High | Continuing pimavanserin after response significantly reduced relapse of psychosis vs switching to placebo (HR 0.35; 95% CI 0.17–0.73). |
| 26 Meltzer et. Al. 2010 | Multicenter | 60 | 46 | 14 | 70.9 | 2 | Presence of visual and/or auditory hallucinations and/or delusions of **moderate-to-severe frequency and severity for ≥4 weeks**, with **NPI psychosis severity ≥4** (hallucinations + delusions sections). | Pimavanserin | 29 | Placebo | 31 | High | Pimavanserin improved several psychosis measures (SAPS global hallucinations/delusions and UPDRS-I thought disorder) **without worsening motor function** and with an **AE profile similar to placebo** over 4 weeks. |
| 27 Rabey et. Al. 2007 | Israel | 58 | 33 | 25 | 72 | 3 | Severe visual or auditory hallucinations and/or delusions that significantly affected quality of life (CGI-S ≥4 required). | Quetiapine | 30 | Placebo | 28 | High | In psychotic PD, 12 weeks of quetiapine (mean ~119 mg/day) was well-tolerated but **not superior to placebo** on BPRS/CGI, with high dropout potentially impacting results. |
| 28 Brier et. Al. 2002 | Europe | 83 | 58 | 25 | 71.7 | 1 | **Treatment-associated psychosis per DSM-IV**, with hallucinations or delusions in the prior 2 weeks and **NPI Hallucinations or Delusions item ≥2 at both screening and randomization**. | Olanzapine | 90 | Placebo | 70 | High | **Olanzapine did not demonstrate superior antipsychotic efficacy versus placebo and was associated with significant worsening on UPDRS total, Motor, and ADL scores in both trials.** |
| 29 Cummings et. Al. 2014 | USA | 199 | 116 | 69 | 72.6 | 1.5 | Adults with PD who met PDP diagnostic criteria with hallucinations and/or delusions present ≥1 month, occurring at least weekly prior to screening, severe enough to warrant antipsychotic treatment, and meeting SAPS/SAPS-PD threshold at baseline. | Pimavanserin | 105 | Placebo | 94 | High | Pimavanserin significantly improved psychosis (SAPS-PD) versus placebo over 6 weeks **without worsening motor function**. |
| 30 The French Clozapine Parkinson Study Group 1999 | France | 60 | 30 | 30 | 72 | 1 | **Persistent psychosis despite stopping anticholinergics, amantadine, and selegiline, and no improvement (or unacceptable motor worsening) when trying to withdraw dopamine agonists or reduce levodopa.** | Clozapine | 32 | Placebo | 28 | High | **Low-dose clozapine (≤50 mg/day) over 4 weeks significantly improved psychosis in PD (CGI and PANSS-positive) without overall motor worsening and with acceptable tolerability (notably somnolence and some transient parkinsonism).** |
| 31 Mogante et. Al. 2002 | Italy | 20 | 10 | 10 | 68.5 | 3 | DSM-IV drug-induced psychosis (from antiparkinsonian drugs) with ≥4 weeks’ history and BPRS hallucinations and/or delusions item score ≥3 at baseline. | Quetiapine | 10 | Clozapine | 10 | High | Both quetiapine and clozapine significantly improved psychosis (BPRS, CGI-S) with no between-group difference; clozapine modestly improved motor function (UPDRS III), while quetiapine was largely unchanged (with slight worsening at higher doses). |
| 32 Pollak et. Al. 2004 | France | 60 | 32 | 28 | 71.95 | 5 | Idiopathic PD patients with **hallucinations and/or delusions** rated **PANSS P1 or P3 ≥ 4 for ≥ 2 weeks**, after standard attempts to reduce dopaminergic therapy; MMSE ≥ 20. | Clozapine | 32 | Placebo | 28 | High | **Low-dose clozapine (≤ 50 mg/day) significantly improved PD psychosis without clinically meaningful motor worsening; the benefit dissipated when clozapine was stopped.** |
| 33 Pintor et. Al. 2012 | Spain | 16 | 6 | 12 | 72 | 1 | Psychotic episode induced by substances or secondary to PD per DSM-IV criteria, persisting after dopaminergic dose adjustments and after ≥5-day washout of anticholinergics, amantadine, COMT inhibitors, or selegiline. | Ziprasidone | 8 | Clozapine | 8 | High | Ziprasidone (20–80 mg/day) was at least as effective as low-dose clozapine over 4 weeks for PD psychosis and did not worsen motor symptoms. |
| 34 Goetz et. Al. 2000 | USA | 15 | 7 | 8 | 72.1 | 2 | DSM-IV hallucinations/psychosis occurring at least weekly for ≥30 days. | Olanzapine | 7 | Clozapine | 8 | High | Compared with clozapine, olanzapine aggravated parkinsonism and did not improve psychosis, so it should not be routinely used for hallucinations in PD. |
| 35 Shotbolt et. Al. 2009 | UK | 24 | 16 | 8 | 71.8 | 3 | Hallucinations, suspiciousness, or unusual thought content (delusions) rated >3/7 on the BPRS for >2 weeks. | Quetiapine | 11 | Placebo | 13 | High | Quetiapine (up to 150 mg/day) did not significantly improve psychosis versus placebo and did not worsen motor function, though more patients on quetiapine dropped out (difference not significant). |

Table S2. Ranking table for all treatments for BPRS

|  | **Clozapine** | **Olanzapine** | **Placebo** | **Quetiapine** | **Risperidone** |
| --- | --- | --- | --- | --- | --- |
| Clozapine | Clozapine | 9.29 (-10.08, 28.87) | 10 (-7.27, 27.21) | 2.89 (-15.8, 21.17) | -5.86 (-33.93, 21.83) |
| Olanzapine | -9.29 (-28.87, 10.08) | Olanzapine | 0.69 (-16.19, 17.51) | -6.41 (-26.8, 13.5) | -15.15 (-49.36, 18.68) |
| Placebo | -10 (-27.21, 7.27) | -0.69 (-17.51, 16.19) | Placebo | -7.12 (-19.98, 5.43) | -15.82 (-48.9, 16.9) |
| Quetiapine | -2.89 (-21.17, 15.8) | 6.41 (-13.5, 26.8) | 7.12 (-5.43, 19.98) | Quetiapine | -8.73 (-42.19, 24.67) |
| Risperidone | 5.86 (-21.83, 33.93) | 15.15 (-18.68, 49.36) | 15.82 (-16.9, 48.9) | 8.73 (-24.67, 42.19) | Risperidone |

Figure S1. Ranking of all treatment for BPRS

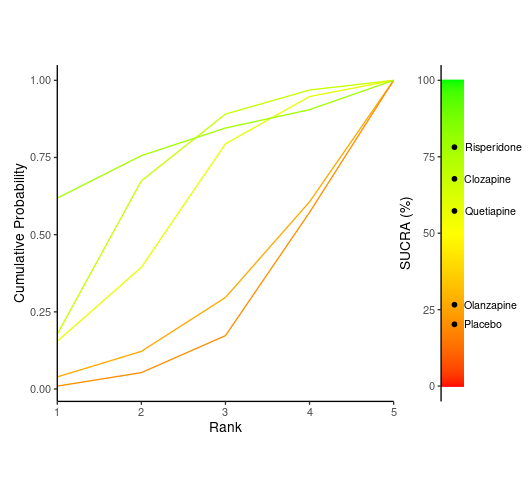

Table S3. Ranking table for all treatments for CGI-S

|  | **Clozapine** | **Olanzapine** | **Pimavanserin** | **Placebo** | **Quetiapine** | **Risperidone** | **Ziprasidone** |
| --- | --- | --- | --- | --- | --- | --- | --- |
| Clozapine | Clozapine | -0.69 (-4.35, 3) | 1.62 (-1.38, 4.66) | 1.21 (-1.39, 3.81) | 0.12 (-3.6, 3.92) | -0.82 (-13.62, 11.42) | -0.2 (-3.29, 2.89) |
| Olanzapine | 0.69 (-3, 4.35) | Olanzapine | 2.33 (-0.71, 5.32) | 1.9 (-0.72, 4.49) | 0.82 (-2.95, 4.59) | -0.1 (-13.41, 12.57) | 0.5 (-4.3, 5.26) |
| Pimavanserin | -1.62 (-4.66, 1.38) | -2.33 (-5.32, 0.71) | Pimavanserin | -0.43 (-1.94, 1.09) | -1.51 (-4.64, 1.64) | -2.42 (-15.55, 10.16) | -1.83 (-6.15, 2.44) |
| Placebo | -1.21 (-3.81, 1.39) | -1.9 (-4.49, 0.72) | 0.43 (-1.09, 1.94) | Placebo | -1.08 (-3.8, 1.66) | -2.01 (-15.06, 10.49) | -1.41 (-5.44, 2.6) |
| Quetiapine | -0.12 (-3.92, 3.6) | -0.82 (-4.59, 2.95) | 1.51 (-1.64, 4.64) | 1.08 (-1.66, 3.8) | Quetiapine | -0.94 (-14.21, 11.8) | -0.32 (-5.22, 4.5) |
| Risperidone | 0.82 (-11.42, 13.62) | 0.1 (-12.57, 13.41) | 2.42 (-10.16, 15.55) | 2.01 (-10.49, 15.06) | 0.94 (-11.8, 14.21) | Risperidone | 0.63 (-12.01, 13.66) |
| Ziprasidone | 0.2 (-2.89, 3.29) | -0.5 (-5.26, 4.3) | 1.83 (-2.44, 6.15) | 1.41 (-2.6, 5.44) | 0.32 (-4.5, 5.22) | -0.63 (-13.66, 12.01) | Ziprasidone |

Figure S2. Ranking of all treatment pairs for CGI-S

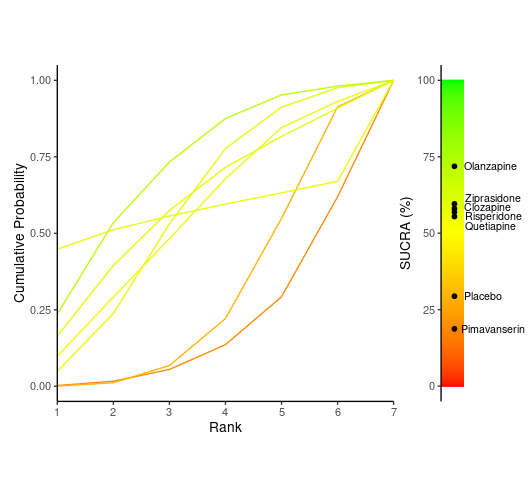

Table S4. Ranking for all treatments in UPDRS-II

|  | **Clozapine** | **Olanzapine** | **Pimavanserin** | **Placebo** | **Quetiapine** | **Risperidone** | **Ziprasidone** |
| --- | --- | --- | --- | --- | --- | --- | --- |
| Clozapine | Clozapine | 0.51 (-7, 7.05) | 0.99 (-5.61, 7.48) | 0.82 (-2.96, 4.57) | 0.29 (-3.95, 4.15) | 4.19 (-5.12, 13.63) | -3.07 (-16.36, 10.46) |
| Olanzapine | -0.51 (-7.05, 7) | Olanzapine | 0.46 (-6.99, 8.93) | 0.31 (-5.14, 6.76) | -0.23 (-6.64, 6.9) | 3.71 (-7.6, 15.87) | -3.49 (-18.29, 11.92) |
| Pimavanserin | -0.99 (-7.48, 5.61) | -0.46 (-8.93, 6.99) | Pimavanserin | -0.16 (-5.52, 5.17) | -0.69 (-7.12, 5.33) | 3.2 (-8.11, 14.62) | -4.05 (-18.76, 10.92) |
| Placebo | -0.82 (-4.57, 2.96) | -0.31 (-6.76, 5.14) | 0.16 (-5.17, 5.52) | Placebo | -0.52 (-3.94, 2.5) | 3.37 (-6.65, 13.47) | -3.88 (-17.69, 10.14) |
| Quetiapine | -0.29 (-4.15, 3.95) | 0.23 (-6.9, 6.64) | 0.69 (-5.33, 7.12) | 0.52 (-2.5, 3.94) | Quetiapine | 3.92 (-6.12, 14.23) | -3.31 (-17.17, 10.85) |
| Risperidone | -4.19 (-13.63, 5.12) | -3.71 (-15.87, 7.6) | -3.2 (-14.62, 8.11) | -3.37 (-13.47, 6.65) | -3.92 (-14.23, 6.12) | Risperidone | -7.25 (-23.51, 9.06) |
| Ziprasidone | 3.07 (-10.46, 16.36) | 3.49 (-11.92, 18.29) | 4.05 (-10.92, 18.76) | 3.88 (-10.14, 17.69) | 3.31 (-10.85, 17.17) | 7.25 (-9.06, 23.51) | Ziprasidone |

Figure S3. Ranking of all treatment pairs in UPDRS-II

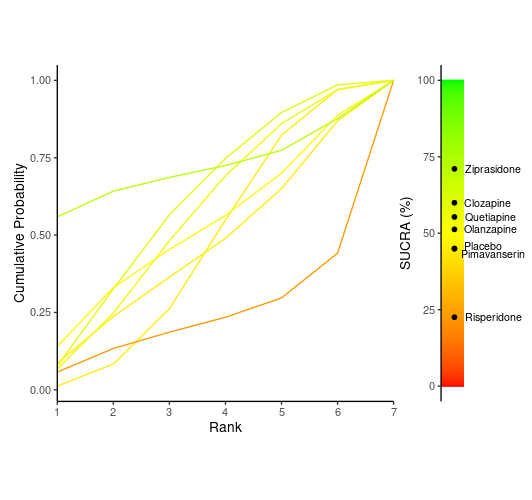

Figure S4. Risk of Bias

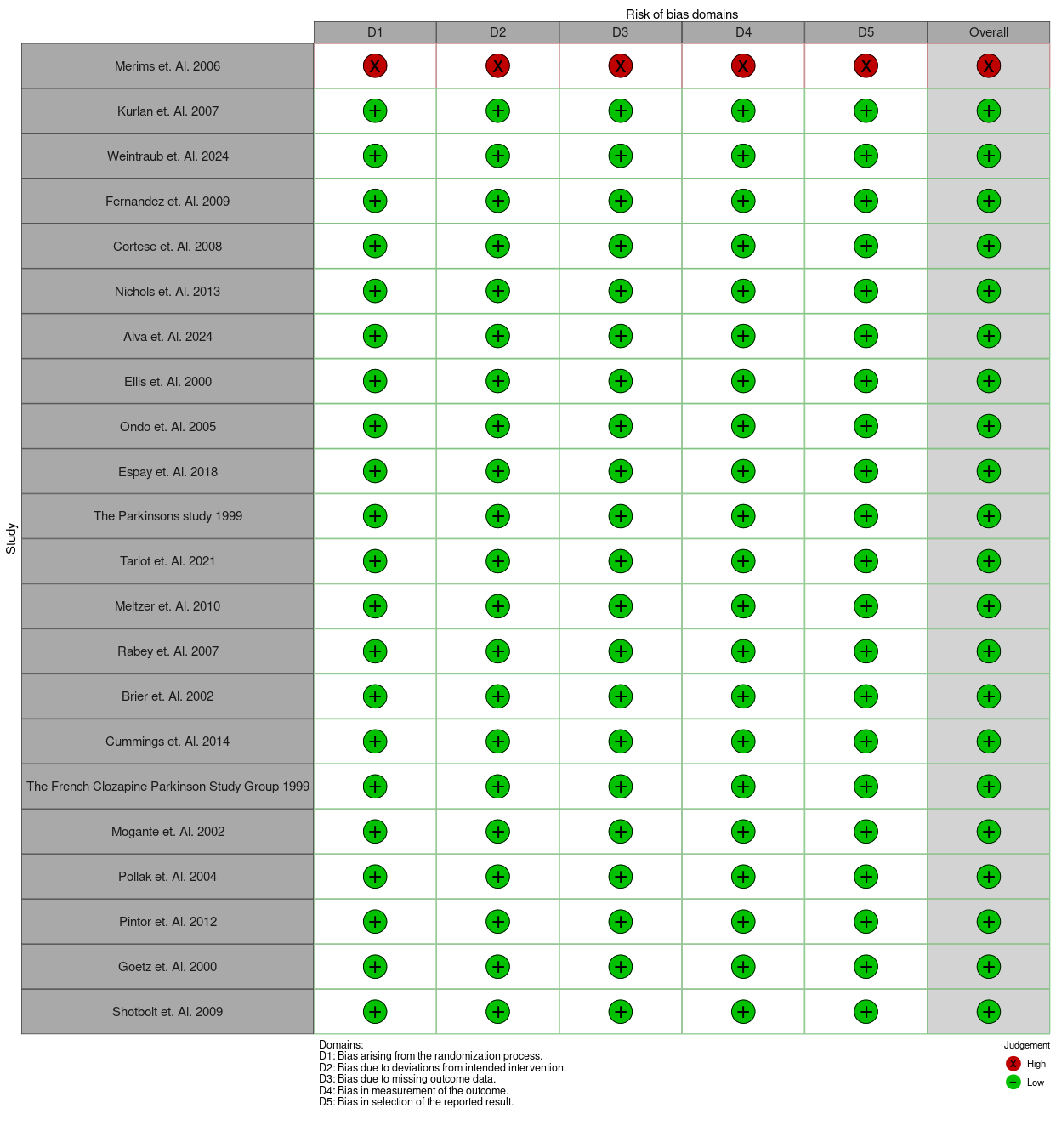
